# Gaps and Opportunities for Data Systems and Economics to Support Priority Setting for Climate-Sensitive Infectious Diseases in Sub-Saharan Africa: A Rapid Scoping Review

**DOI:** 10.1101/2024.09.20.24314043

**Authors:** Ellie A. Delight, Ariel A. Brunn, Francis Ruiz, Jessica Gerard, Jane Falconer, Yang Liu, Bubacarr Bah, Bernard Bett, Benjamin Uzochukwu, Oladeji K. Oloko, Esther Njuguna, Kris A. Murray

## Abstract

Climate change alters risks associated with climate-sensitive infectious diseases (CSIDs) with pandemic potential. This poses additional threats to already vulnerable populations, further amplified by intersecting social factors, such as gender and socioeconomic status. Currently, critical evidence gaps and inadequate institutional and governance mechanisms impact on the ability for African States to prevent, detect and respond to CSIDs. The aim of this study was to explore the role of data systems and economics to support priority setting for CSID preparedness in sub-Saharan Africa.

We conducted a rapid scoping review to identify existing knowledge and gaps relevant to economics and data systems. A literature search was performed across six bibliographic databases in November 2023. A list of 14 target pathogens, identified by the World Health Organization as Public Health Emergencies of International Concern or R&D Blueprint Pathogens, was adopted and compared to a database of CSIDs to determine relevant inclusion criteria. Extracted data were synthesised using bibliometric analysis, thematic topic categorisation, and narrative synthesis to identify research needs, evidence gaps, and opportunities for priority setting.

We identified 68 relevant studies. While African author involvement has been increasing since 2010, few studies were led by senior authors from African institutions. Data system studies (n = 50) showed broad coverage across CSIDs and the WHO AFRO region but also a high degree of heterogeneity, indicating a lack of clearly defined standards for data systems related to pandemic preparedness. Economic studies (n = 18) primarily focused on COVID-19 and Ebola and mostly originated from South Africa. Both data system and economic studies identified limited data sharing across sectors and showed a notable absence of gender sensitivity analyses. These significant gaps highlight important opportunities to support priority setting and decision-making for pandemic preparedness, ultimately leading to more equitable health outcomes.

## Introduction

Rising greenhouse gas (GHG) emissions impact the frequency and intensity of climate hazards (e.g., temperature, rainfall) and extremes (e.g., droughts, floods, and heatwaves), which can aggravate the transmission of climate-sensitive infectious diseases (CSIDs) (1). Additionally, changing climate patterns alter ecological niches where humans, animals and vectors are able to survive (2). In conjunction with changing land-use patterns, these altered ecological settings and climate hazards can impact pathogen exchange among previously isolated vectors, wildlife, livestock, and human populations (3, 4). This can expose immunologically naïve populations to CSIDs, leading to increased risk of CSID pandemics. Notably, all eight of the viral diseases declared as Public Health Emergencies of International Concern (PHEIC) by the World Health Organization (WHO) (5) are climate-sensitive according to a recent review (1), including COVID-19 and Mpox. Additionally, eight of the nine pathogens prioritised by the WHO Research & Development (R&D) Blueprint list are classed as climate-sensitive (6). The R&D list helps to prioritise R&D resources towards pathogens that pose significant public health risk due to their epidemic potential or insufficient control measures.

Vulnerability to CSIDs is further driven by marginalisation coupled with social determinants of health such as gender, socioeconomic status, race, and disability. The intersectional gendered risks of CSIDs were evident in the COVID-19 pandemic, where the case fatality rate was higher among men (7), but women faced greater socioeconomic impacts and increased risk of domestic violence as a result of lockdown restrictions (8, 9). Adopting an intersectional gender lens in CSID research can enable a more contextualised and thorough understanding of the aetiology of CSIDs, advocating for an equitable approach to CSID pandemic response tailored to men, women, boys, girls, and other gender identities (10).

A growing body of evidence demonstrates the disproportionate impact of climate change across countries in sub-Saharan Africa, despite minimal contributions to GHG emissions (11–13). Some limited studies have evaluated the risk of CSIDs in sub-Saharan Africa; for instance, in low-income, rural communities in the Sahel region of West Africa, daily high temperatures (above 41.1 °C compared to 36.4°C median) and low rainfall (below 10mm compared to 14mm median) were associated with increased deaths from CSIDs (13). Additionally, climate hazards such as flooding can prevent access to healthcare services, which can cause diagnostic and treatment delays (14), impeding early detection of outbreaks and raising the risk of uncontrolled disease spread and pandemic emergence.

The ability to prepare for the outsized risks of CSID pandemics in sub-Saharan Africa is limited by weak institutional collaborations or integrated governance mechanisms. This hinders data-driven decision-making and appropriate and judicious allocation of scarce resources. Successful policy response requires capacity on both sides of the research-policy nexus, including skilled researchers generating robust data and relevant economic evaluation, and presence of formalised systems or bodies to build confidence and public support for the deployment of public funds during a public health crisis. Interoperable data systems that provide near real-time information across human, animal, and environmental domains inform decision-making for the detection and prevention of CSIDs. Additionally, data systems can help to inform decision-making across later stages of pandemic preparedness; for example by helping to tailor control strategies to regional climate differences that would otherwise hamper response strategies (15). Priority setting is a process that can help manage complex decision-making and may be especially beneficial in pandemic response, particularly in low-resource settings where policy makers must balance emergency needs with regular health programs. Recent outbreaks, such as the 2014 Ebola outbreak in West Africa (16) and more recently with COVID-19 globally, illustrate the long-lasting economic and health consequences of pandemics. Understanding the cost-effectiveness of interventions and response can aid decision-makers in allocating scarce resources during pandemic response. This can incorporate economic evaluation (such as cost-effectiveness analyses (CEA), cost-utility analyses (CUA), or cost-benefit analyses (CBA)), which are ideally institutionalised and include approaches such as Health Technology Assessment (HTA) that consider a body of evidence and multi-stakeholder perspectives.

Effective priority setting with respect to pandemic preparedness and CSIDs, or indeed in any situation where a resource allocation decision needs to be made, should be informed by evidence of both likely benefits and cost-effectiveness (17). In turn, such assessments of value depend on credible sources of information, even though assumptions and the need to make qualitative judgements are unavoidable. Priority setting is therefore facilitated by having fit-for-purpose (and cost-effective) data systems that capture contextualised information on epidemiology, vulnerable groups, resource use, and other relevant socio-economic characteristics. As such, data systems are key to informing CSID control strategies, and supporting the parameterisation of economic evaluations that help to consider the benefits of available strategies against the resources required to implement them.

To identify existing knowledge and gaps relevant to data system and economic studies for priority setting of CSIDs with pandemic potential, we extended a conceptual framework across four stages of pandemic preparedness: Prevention (Stage 1): pre-epidemic preparedness; Detection (Stage 2): identify, investigate, evaluate risk; Response (Stage 3): outbreak response & containment; Evaluation (Stage 4): Post-epidemic evaluation (18) (Fig 1).

**Fig 1.**
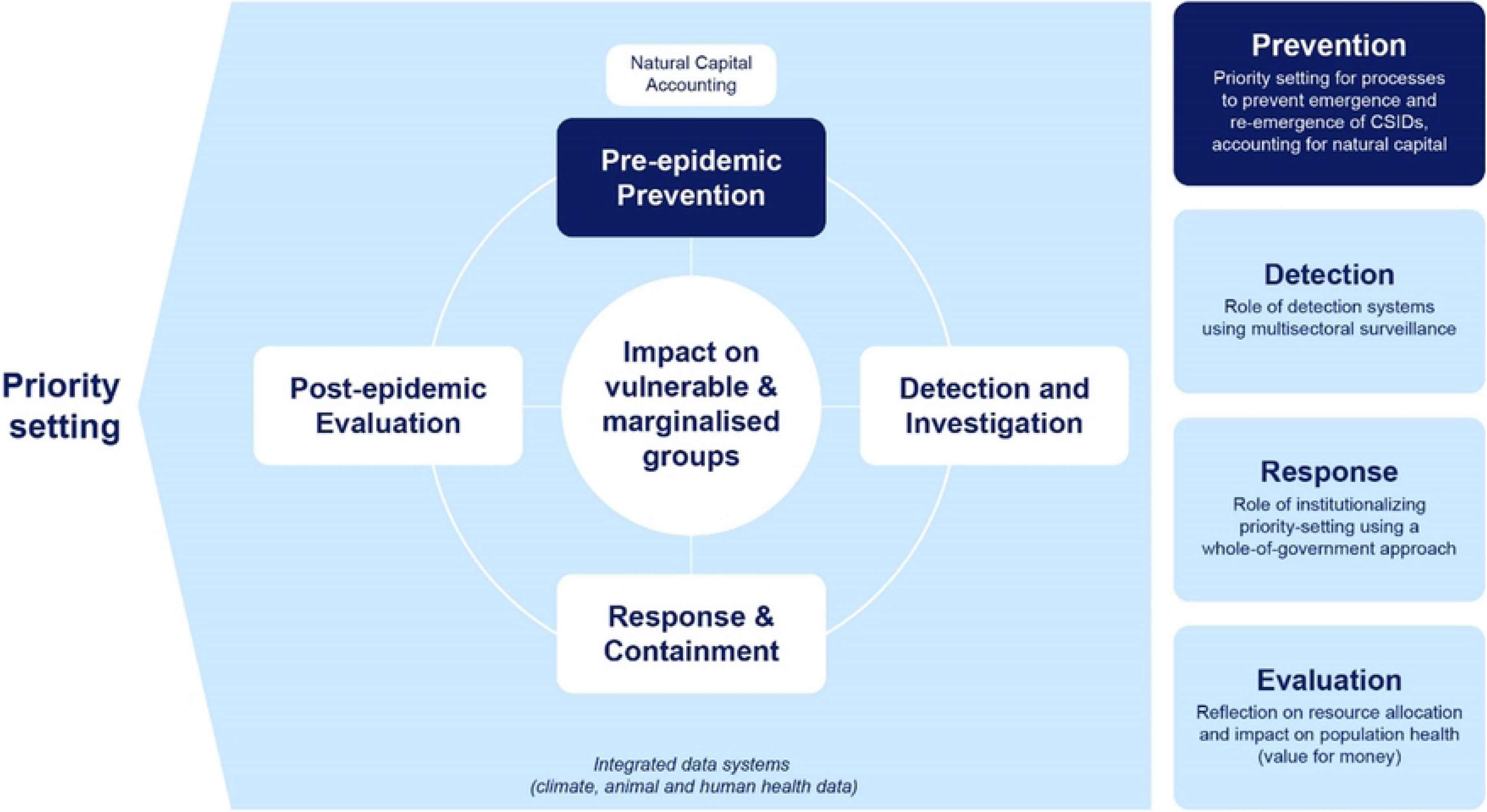
Pandemic Preparedness Framework for Climate Sensitive Infectious Disease in Africa. Adapted from: World Health Organization. (2014). Ebola and Marburg virus disease epidemics: Preparedness, alert, control, and evaluation. https://www.who.int/publications-detail-redirect/WHO-HSE-PED-CED-2014.05

### Rationale

Low- and middle-income countries contribute the least to anthropogenic climate change but are highly vulnerable to its impacts, with important implications for public health. It is crucial to identify evidence gaps, establish future research priorities, and strengthen stakeholder networks at the climate-health intersection to direct research funding and inform policies that enhance health outcomes and resilience of health systems for vulnerable populations. This necessitates multidisciplinary and comprehensive government approaches, leveraging more integrative disciplines such as One Health and Planetary Health, with an emphasis on intersectional gendered analysis to ensure representation of marginalised communities. By deepening our understanding of the climate-health nexus, we can develop more effective strategies for preventing, detecting, and responding to disease outbreaks.

### Objectives

i. To map key themes and entry points for future African-led research and to identify gaps in evidence for priority setting using economic evaluation and data systems for policy development;
ii. To use bibliometric analysis to assess institutional stakeholder networks and to evaluate trends in research development on key themes and outputs included in the review;
iii. To identify applications for a transdisciplinary and gender-intersectional approach for CSID preparedness and response.

### Research questions

i. What are the key themes, in terms of economic evaluation (including cost-effectiveness) and priority setting, that describe how climate change is incorporated into pandemic preparedness and outbreak response planning?
ii. What data systems are currently in use or needed to support priority setting for integrated and multisectoral epidemic or pandemic planning for climate-sensitive infectious diseases in Africa?
iii. What subnational, national, and regional structures are in place to support joined-up data sharing for CSID outbreak detection and response within and between African countries and the international community?

## Methods

### Rapid Scoping Review

The literature review was conducted as a rapid scoping review. Scoping reviews provide a structured approach to mapping existing knowledge, highlighting common themes, and identifying evidence gaps. Rapid reviews streamline these methods, facilitating quicker knowledge translation, especially when urgent policy or strategy decisions are required (19).

### Protocol and registration

This rapid scoping review was informed by the Preferred Reporting Items for Systematic Reviews and Meta-Analyses for Scoping Reviews (PRISMA-ScR). The protocol for this study is registered at the Open Science Framework (OSF) repository (20) and is available on MedRxiv (21).

### Eligibility criteria

The eligibility criteria are outlined in Table 1.

**Table 1.**
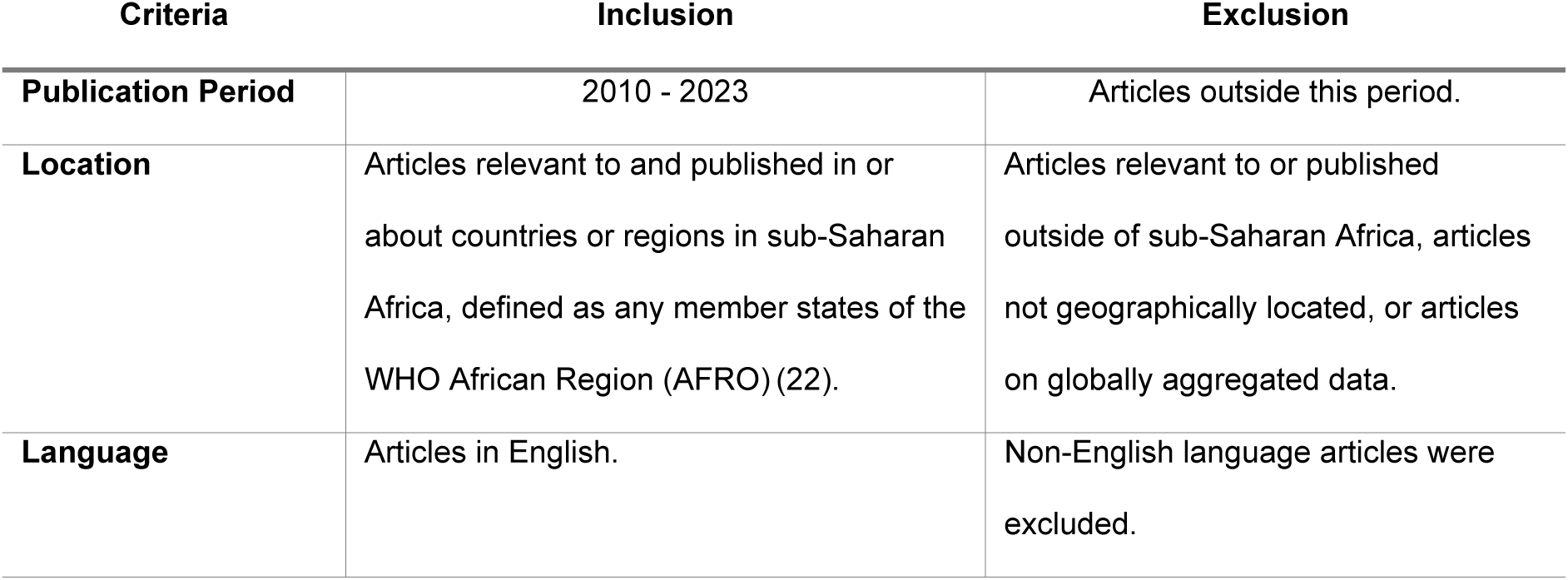
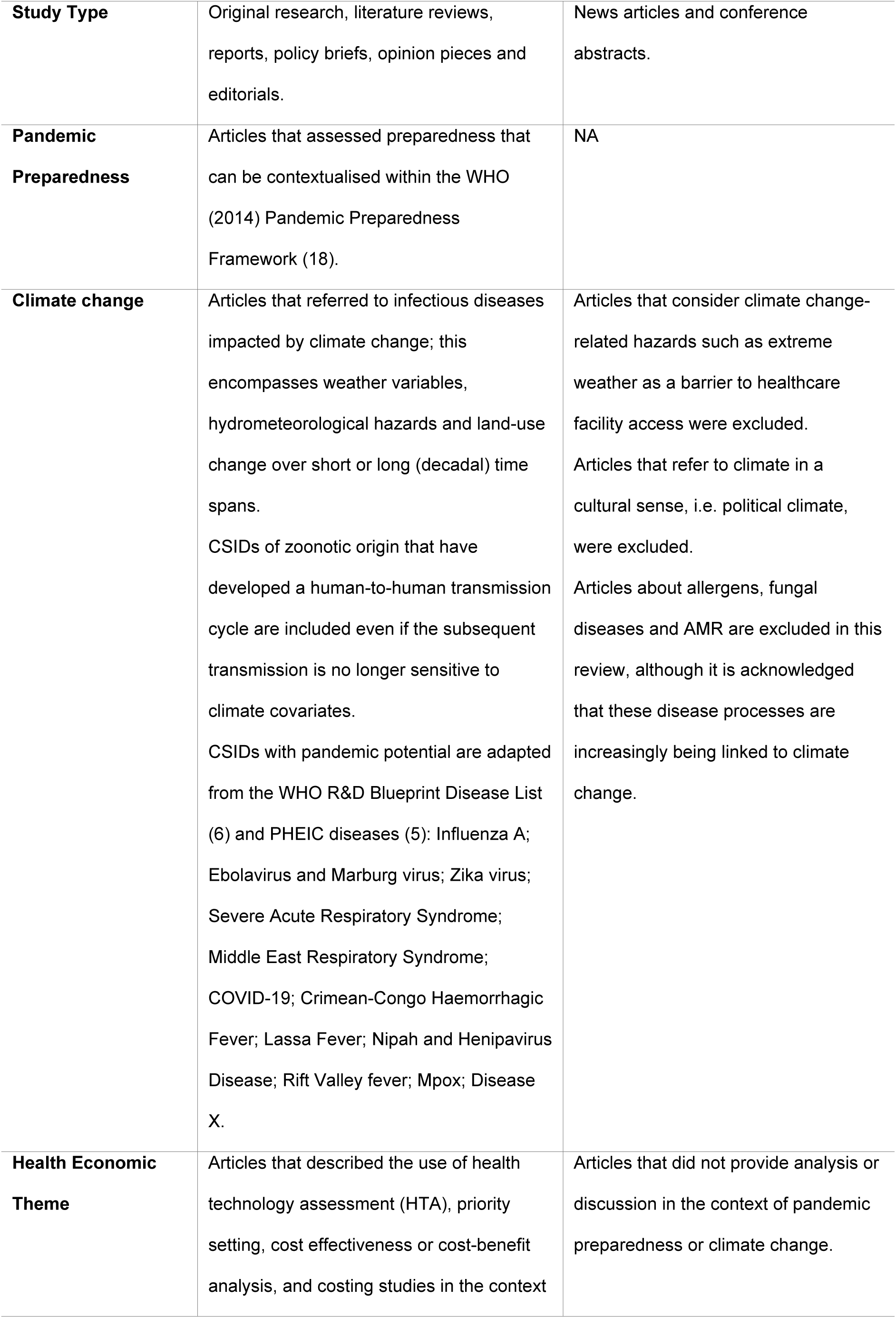
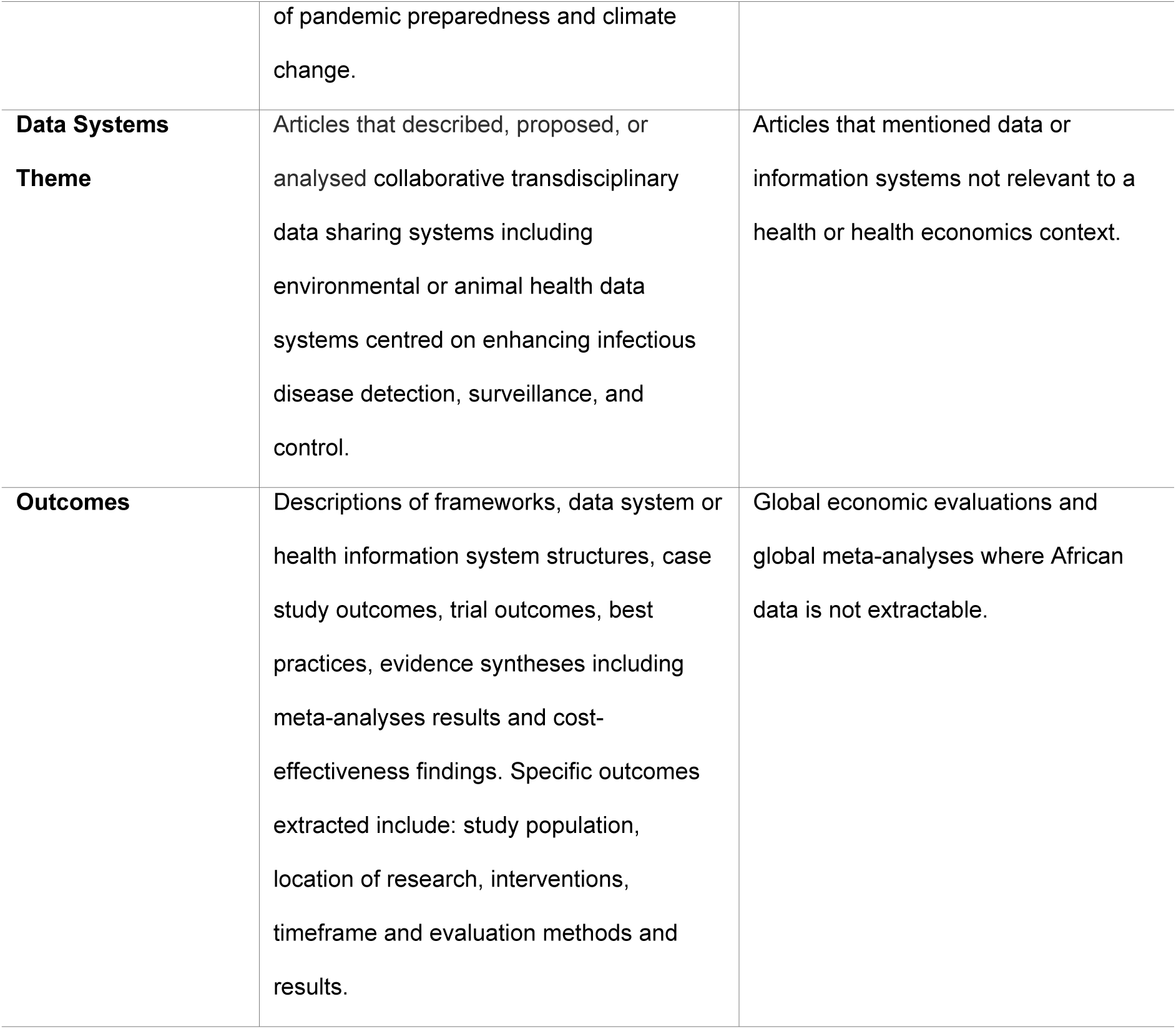
Scoping Review Eligibility Criteria.

| Criteria | Inclusion | Exclusion |
| --- | --- | --- |
| **Publication Period** | 2010 - 2023 | Articles outside this period. |
| **Location** | Articles relevant to and published in or about countries or regions in sub-Saharan Africa, defined as any member states of the WHO African Region (AFRO) (22). | Articles relevant to or published outside of sub-Saharan Africa, articles not geographically located, or articles on globally aggregated data. |
| **Language** | Articles in English. | Non-English language articles were excluded. |
| **Study Type** | Original research, literature reviews, reports, policy briefs, opinion pieces and editorials. | News articles and conference abstracts. |
| **Pandemic Preparedness** | Articles that assessed preparedness that can be contextualised within the WHO (2014) Pandemic Preparedness Framework (18). | NA |
| **Climate change** | Articles that referred to infectious diseases impacted by climate change; this encompasses weather variables, hydrometeorological hazards and land-use change over short or long (decadal) time spans.<br>CSIDs of zoonotic origin that have developed a human-to-human transmission cycle are included even if the subsequent transmission is no longer sensitive to climate covariates.<br>CSIDs with pandemic potential are adapted from the WHO R&D Blueprint Disease List (6) and PHEIC diseases (5): Influenza A; Ebolavirus and Marburg virus; Zika virus; Severe Acute Respiratory Syndrome; Middle East Respiratory Syndrome; COVID-19; Crimean-Congo Haemorrhagic Fever; Lassa Fever; Nipah and Hendra virus; Disease X. | Articles that consider climate change-related hazards such as extreme weather as a barrier to healthcare facility access were excluded.<br>Articles that refer to climate in a cultural sense, i.e. political climate, were excluded.<br>Articles about allergens, fungal diseases and AMR are excluded in this review, although it is acknowledged that these disease processes are increasingly being linked to climate change. |
| **Health Economic Theme** | Articles that described the use of health technology assessment (HTA), priority setting, cost effectiveness or cost-benefit analysis, and costing studies in the context | Articles that did not provide analysis or discussion in the context of pandemic preparedness or climate change. |
|  | of pandemic preparedness and climate change. |  |
| **Data Systems Theme** | Articles that described, proposed, or analysed collaborative transdisciplinary data sharing systems including environmental or animal health data systems centred on enhancing infectious disease detection, surveillance, and control. | Articles that mentioned data or information systems not relevant to a health or health economics context. |
| **Outcomes** | Descriptions of frameworks, data system or health information system structures, case study outcomes, trial outcomes, best practices, evidence syntheses including meta-analyses results and cost-effectiveness findings. Specific outcomes extracted include: study population, location of research, interventions, timeframe and evaluation methods and results. | Global economic evaluations and global meta-analyses where African data is not extractable. |

### Information sources and search

The search strategy was constructed by a library information professional with a focus on four search concepts, namely:

- Pandemic preparedness AND
- Climate change AND
- Economic evaluation (including HTAs and priority setting) OR
- Data systems

The search was conducted in November 2023 across six bibliographic databases: OvidSP Medline, OvidSP Embase, OvidSP Global Health, EBSCOhost Africa-Wide Information, OvidSP Econlit and Clarivate Analytics Web of Science Core Content. Search terms were first tested in one database prior to implementation in the other five. Additional sources of literature from work previously conducted by co-authors were also included. The complete search strategies for all sources is published at the London School of Hygiene & Tropical Medicine Data Compass (23).

### Selection of sources of evidence

Articles were deduplicated and imported into Covidence software (24). Screening was conducted by single independent reviewers through a three-stage process: title, abstract, and full text screening. At each stage, articles were screened using the “Most relevant” option in Covidence, which employs a machine learning algorithm to predict study relevance based on screening of at least 25 studies (25).

Pilot review stages were conducted prior to the abstract and full text screening stages. Prior to the abstract screening stage, the complete team of nine reviewers independently screened abstracts from the same 20 articles and discussed discrepancies to improve consistency. Once consensus was reached, two reviewers independently screened the abstracts for relevance (see S1 Table). Prior to the full text screening stage, two reviewers screened three sets of ten full text publications to iteratively refine screening questions. Once consensus was reached, nine reviewers independently screened the full texts against the full text screening criteria (Fig 2). Any article deemed relevant at the full text screening stage was screened in duplicate by another reviewer to reach consensus.

**Fig 2.**
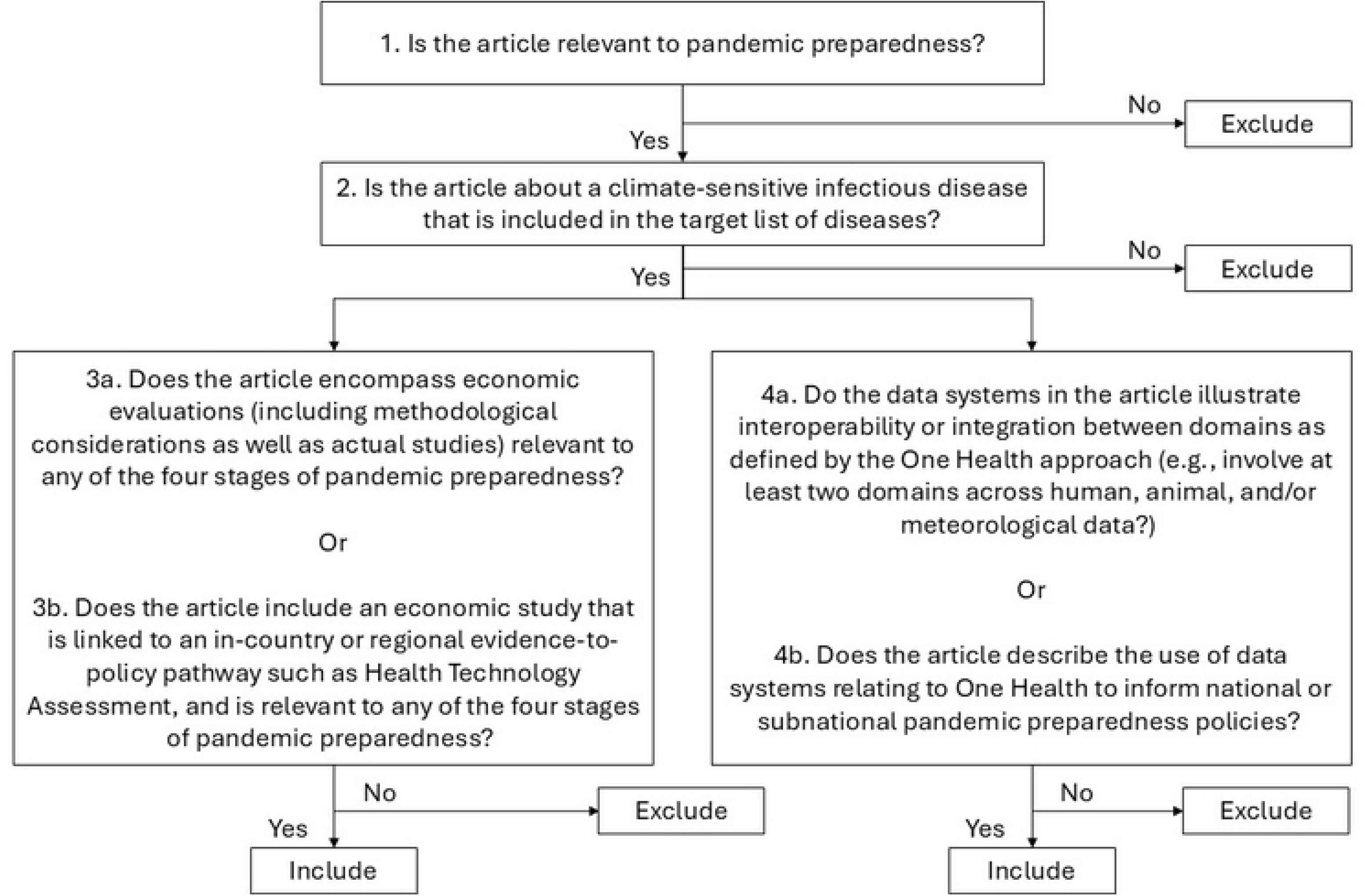
Full text screening criteria.

**Fig 3.**
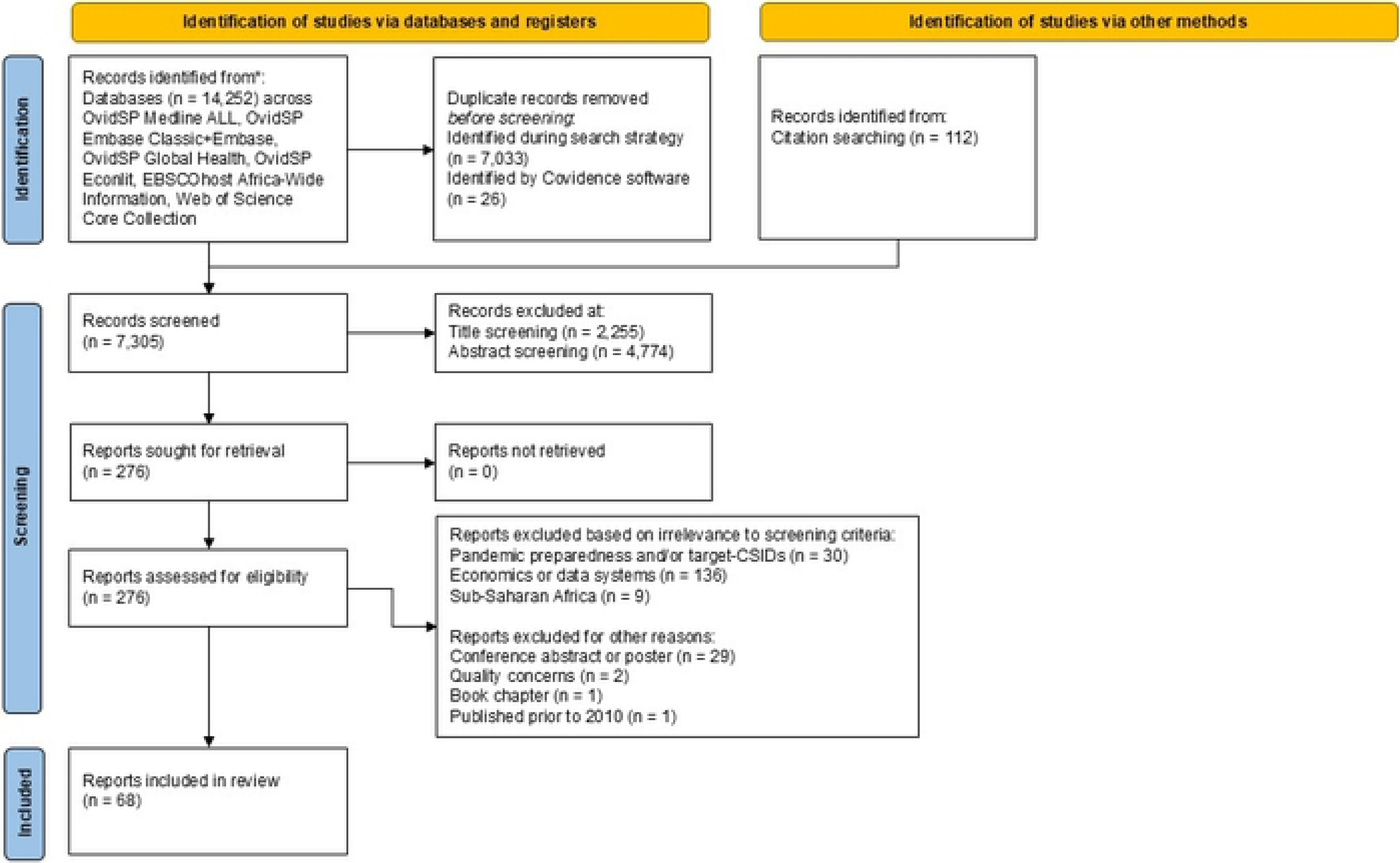
PRISMA-ScR flow diagram of study selection.

### Data charting process

The data charting process was conducted using a data extraction form in Covidence designed by one reviewer and reviewed *a priori* by two reviewers with expertise in the relevant areas. The form was tested against 15 papers by one reviewer and iteratively refined prior to use.

### Data items

The data extracted included article meta-data and thematic content. Article meta-data included year of publication, publication source (institution and country), first and last authors’ names and institutions, collaborating institutions, source of funding, ethical clearance, and type of publication. Topic data were extracted and thematically categorised based on stage of pandemic preparedness and response framework (Fig 1), relevance to target CSIDs (Table 1), and type of data system or economic study. A final mapping of challenges and constraints was conducted by assessing topics with the least volume or least relevant literature, and by conducting a narrative synthesis identifying repeated themes describing evidence gaps arising in the final study set.

### Synthesis of results

The PRISMA flow diagram was created using the PRISMA2020 template (26). We used bibliometric analysis, topic mapping, and narrative summaries to synthesise evidence.

A bibliometric analysis was conducted by assessing the temporal trends of authorship and CSIDs studied as well as geographical distribution of the studies across the WHO AFRO region. Temporal trends of authorship were assessed by categorising the institutional affiliations of all authors as follows: African - all author affiliations located within Africa; African & International - author affiliations located both within Africa and internationally; and International - all author affiliations located outside of Africa.

Topic mapping was conducted by categorising the proportion of data system or economic studies relevant to each stage of pandemic preparedness and each CSID.

A narrative synthesis was conducted by categorising data system and economic studies into sub-categories, summarising identified evidence gaps noted by the study authors, and assessing the gender sensitivity of each study. Data system studies were categorised into two sub-categories: ‘Design’, which included studies that describe the use of multi-disciplinary data systems in pandemic preparedness; and ‘Usage’, which included multidisciplinary analyses that use data from data systems for analyses relevant to pandemic preparedness. The categorisation by this method was chosen to account for the large heterogeneity in the data system studies. Economic studies were categorised into three sub-categories based on study type: Economic evaluations, which include analyses that compare costs of interventions with health outcomes, inclusive of CEAs, CUAs, CBAs, whether or not conducted as part of an HTA; costing studies, which include analyses that consider costs without consideration of health outcomes; and priority setting studies, which are more conceptual or commentary based studies that consider the benefit of economic studies for pandemic preparedness.

The extent that gender was considered across the studies was assessed against definitions provided by the WHO intersectionality gender toolkit for research on infectious diseases of poverty (10), including: Gender-blind research ignores gender norms, roles, and relations; Gender-sensitive research considers inequality generated by unequal gender norms, roles, and relations but takes no remedial action to address it; Gender-specific research considers inequality generated by unequal norms, roles, and relations and takes remedial action to address it but does not change underlying power relations.

All analyses were conducted in Microsoft Excel (27), and figures were produced in R version 4.3.1 via R studio (28). The data extracted from included studies can be found at the OSF (https://osf.io/fn8rw/?view_only=5961c57df32741b884fc2181a0da4ceb).

### Protocol amendments

We applied two protocol amendments to our review. First, we refined the fourth abstract screening question relating to data systems; specifically, we applied more stringent criteria to how we defined data systems. This amendment was required given the large volume of literature utilising data from data systems. Second, we did not conduct a pilot of data extraction in duplicate. Instead, the data extraction form was produced by one reviewer and refined by two reviewers with extensive experience in either economics or data systems. The form was then applied to ten studies by an independent reviewer and iteratively refined prior to use across the final studies.

## Results

### Selection of sources of evidence

The screening process overview is presented in a PRISMA flow chart (Fig 5). A search across six databases resulted in the identification of 14,252 items and an additional 112 studies were retrieved from previous reviews conducted by the research team. After removal of 7,059 duplicate studies, 7,305 studies were screened. During screening, 2,255 studies and then 4,774 studies were removed at the title and abstract screening stages respectively. A total of 276 full texts were screened, culminating in 68 studies that were relevant for data extraction. Of the 68 studies, 50 and 18 were categorised as data system and economic studies, respectively.

### Bibliometric Analysis

#### Temporal publication trends

The frequency of publications increased between 2010 and 2023 (Fig 4a). Authorship trends indicate increasing involvement of researchers based in African institutions in CSID research since 2010, though mixed institutional research teams continue to be dominated by institutions based worldwide up to the most recent year. African-only authorship, where all authors were affiliated with African-based institutions, emerged in 2018 and peaked in 2020. Further breakdown of authorship found that most first authors were affiliated with global institutions (69%) rather than African institutions (29%), and one author was affiliated to both global and African institutions. Similarly, most senior authors were affiliated with institutions outside Africa (63%) compared with African institutions (34%), and two authors had more than one institutional affiliation in and outside of Africa. COVID-19, Ebola, Influenza, and Rift Valley fever were the most studied target CSIDs (Fig 4b).

**Fig 4.**
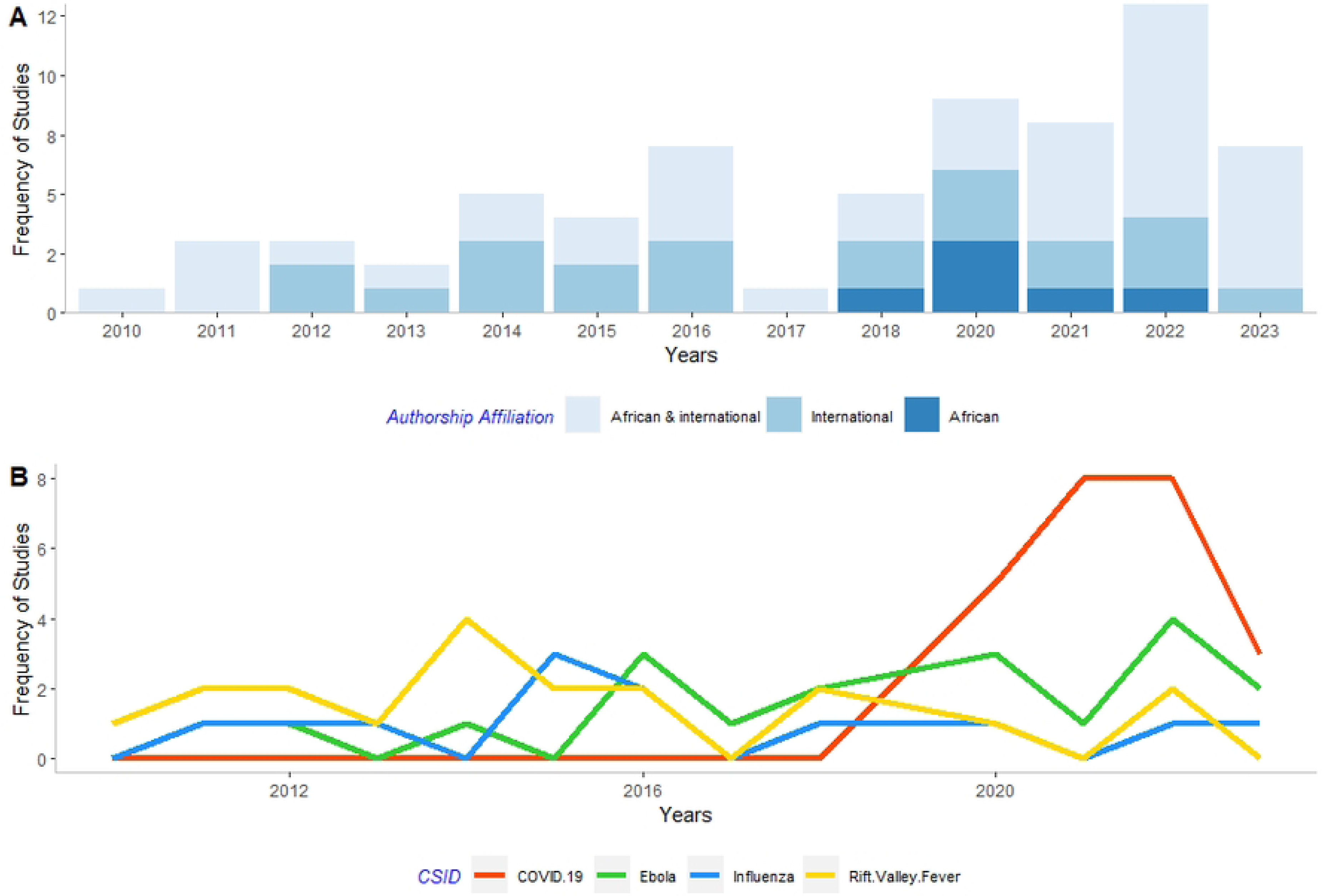
Number of studies published over time, by geographical distribution of authors (A), and four most frequently cited CSIDs (B)

#### Geographical publication trends

Data system studies showed broad coverage across the WHO AFRO region, whereas economic studies were much more concentrated in certain countries (Fig 5). Of the data system studies (n = 50), all countries in the WHO AFRO region were represented in at least one study, largely due to three regional-based studies that included all countries. Nigeria was the most represented country in data system studies, appearing in 36% (18/50) of studies, followed by Ghana and Kenya, which were each represented in 32% (16/50) of studies. In contrast, of the economic studies (n = 18), nearly half of the WHO AFRO countries were not represented at all. South Africa was the most frequently studied country in economic studies, appearing in 28% (5/18) of studies, followed by Ghana and Sierra Leone, each represented in 22% (4/18) of studies.

**Fig 5.**
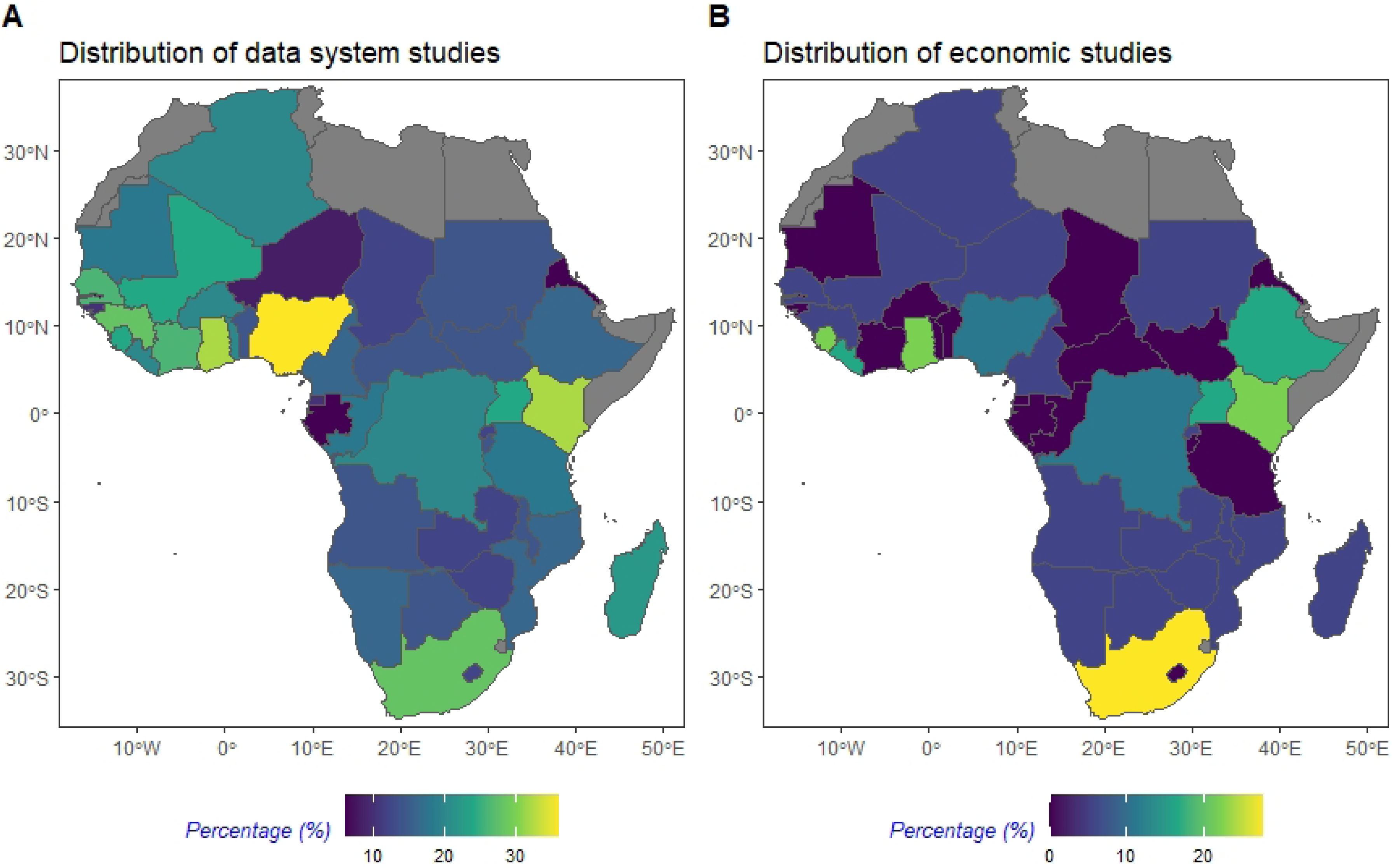
Distribution of (A) data system studies and (B) economic studies. The distribution of countries was plotted as percentage of frequency of country included across data system or economic studies by total number of data system or economic studies. Countries coloured in grey correspond to countries outside of the WHO AFRO region.

### Topic mapping

#### Stage of pandemic preparedness

Topic mapping of studies to pandemic preparedness stages revealed distinct focus areas between data system and economic studies, with data systems being more prevalent in earlier stages of pandemic preparedness while economic studies were more concentrated in the later stages (Fig 6). A higher proportion of data system studies were linked to stages 1 and 2 (46% and 28%, respectively) compared with stages 3 or 4 (19% and 9%, respectively). Conversely, a higher proportion of economic studies were linked to stages 3 or 4 (55% and 26%, respectively) compared with stages 1 or 2 (10% across both stages).

**Fig 6.**
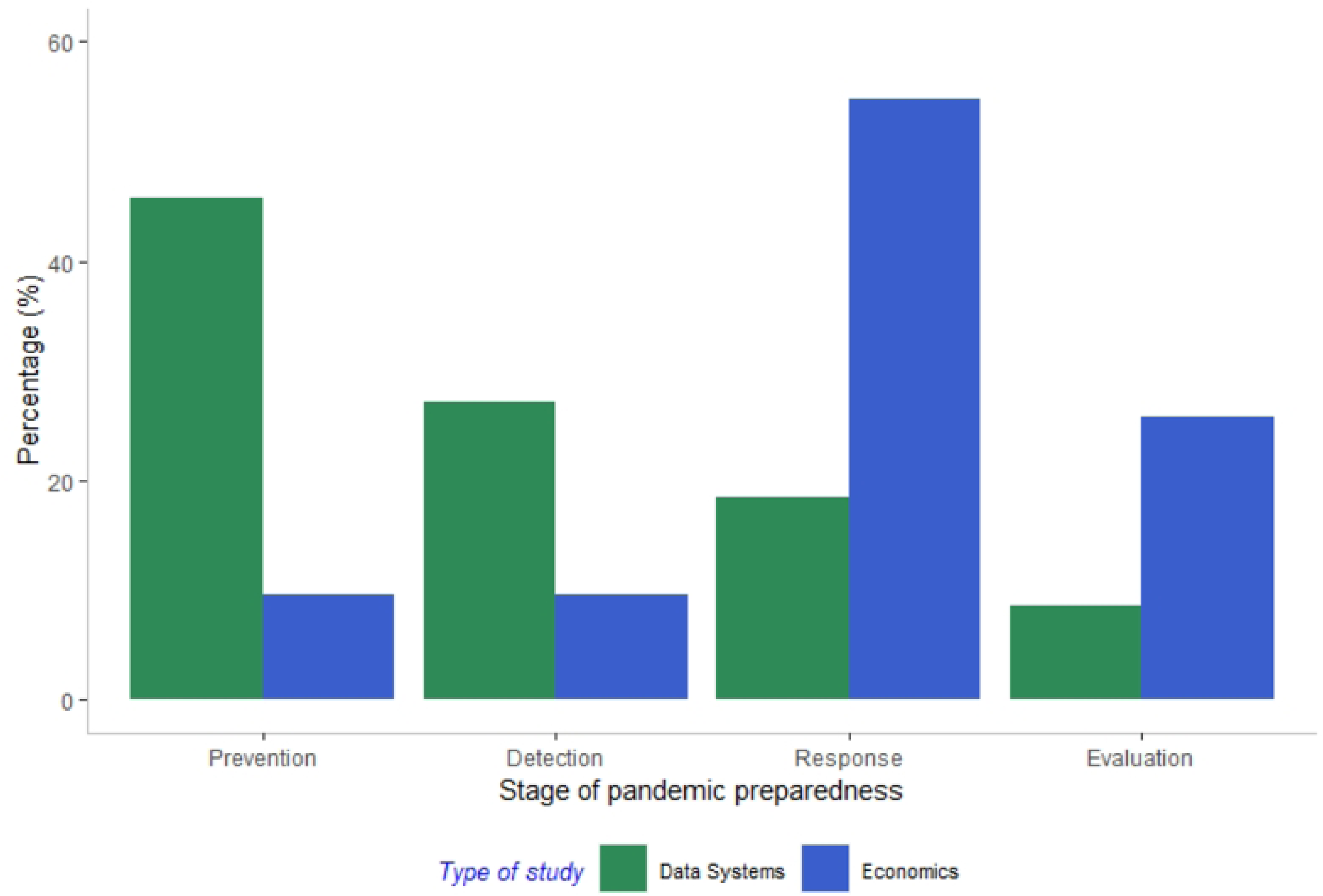
Percentage of data system or economic studies mapped to stages of pandemic preparedness. Studies could be relevant to all four stages; percentage was calculated as tagged stage by total of tagged stages in data system studies (n = 50) or economic studies (n = 18).

#### Climate-sensitive infectious diseases

Topic mapping of studies to CSIDs revealed broad coverage across all CSIDs in data system studies, with Rift Valley fever and Ebola virus disease being the most common (Fig 7). Specifically, data system studies were predominantly focused on five CSIDs: Rift Valley fever (25%), Ebola virus disease (16%), Influenza (14%), COVID-19 (14%), and Lassa fever (9%), although each CSID was represented at least once in the included studies. In contrast, economic studies were restricted to four CSIDs, predominantly focusing on COVID-19 (64%) and Ebola (27%). Influenza and Rift Valley fever were each addressed in 4% of the studies, with both being analysed within the same publication that also covered Ebola (29).

**Fig 7.**
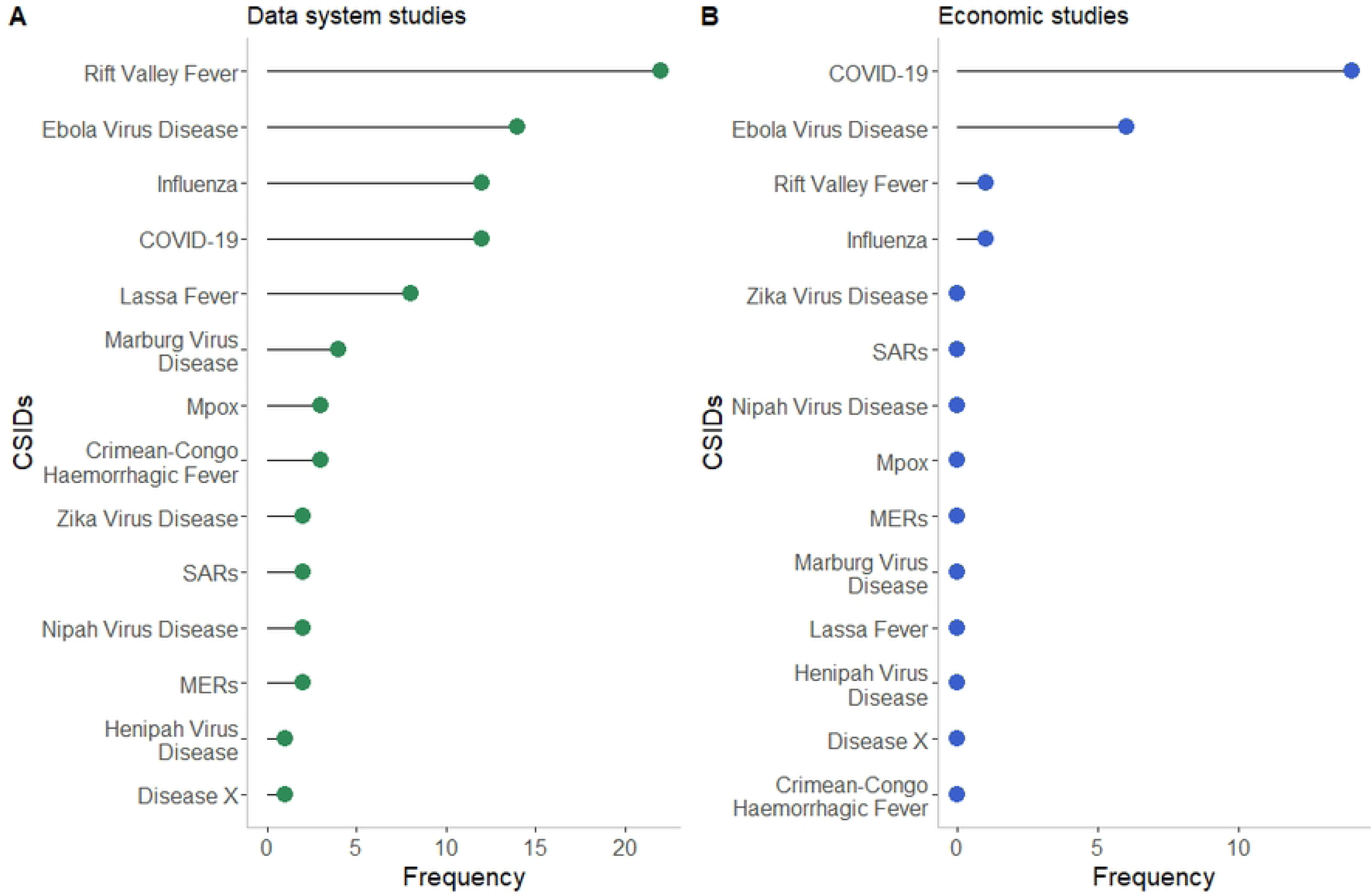
Frequency of CSIDs across (A) data system and (B) economic studies. Studies could be mapped to more than one CSID. Abbreviations: COVID-19: Coronavirus Disease 2019; SARS: Severe Acute Respiratory Syndrome; MERs: Middle East Respiratory Syndrome.

### Narrative Synthesis: Data system studies

The following sections provide a narrative summary mapped to the type of study identified through our screening criteria and the evidence gaps identified by the study authors.

The 50 data system studies were categorised into ‘Design’ studies (n = 19) or ‘Usage’ studies (n = 33). Design studies described data systems in pandemic preparedness, while Usage studies were multidisciplinary analyses that integrated data from data systems across two or more data domains across animal, human, or environmental domains. Three studies were categorised as both Design and Usage studies (30–32).

#### Design data system studies

Of the 19 studies that described data systems in pandemic preparedness (i.e., Design studies), 10 were original research (30, 32–40), five were literature reviews, (41–45), two were outbreak response case studies (46, 47), one was a commentary (48), and one was a letter to the editor (49).

Across the 19 Design data system studies, 28 data systems were described with international, regional, national, or local scales (Fig 8, S2 Table). Seven of the data systems were surveillance systems integrating human, animal, or environmental domains (33, 41–44, 47); six were early warning systems (32, 35, 38, 40, 45); four described community-based surveillance systems (33, 36, 44, 49); two were mobile surveillance tools used in participatory disease surveillance methods (33, 34); two were communication tools designed to share risk maps (30, 45); two were diagnostic laboratory systems (42, 46); two were online genomic sequencing systems (31, 48); two were capacity systems to assess one health capacity of countries (39) or coordinate a pandemic response (46); and one study focused on off-line software designed to aid users to predict the causative pathogen of an outbreak (37).

**Fig 8.**
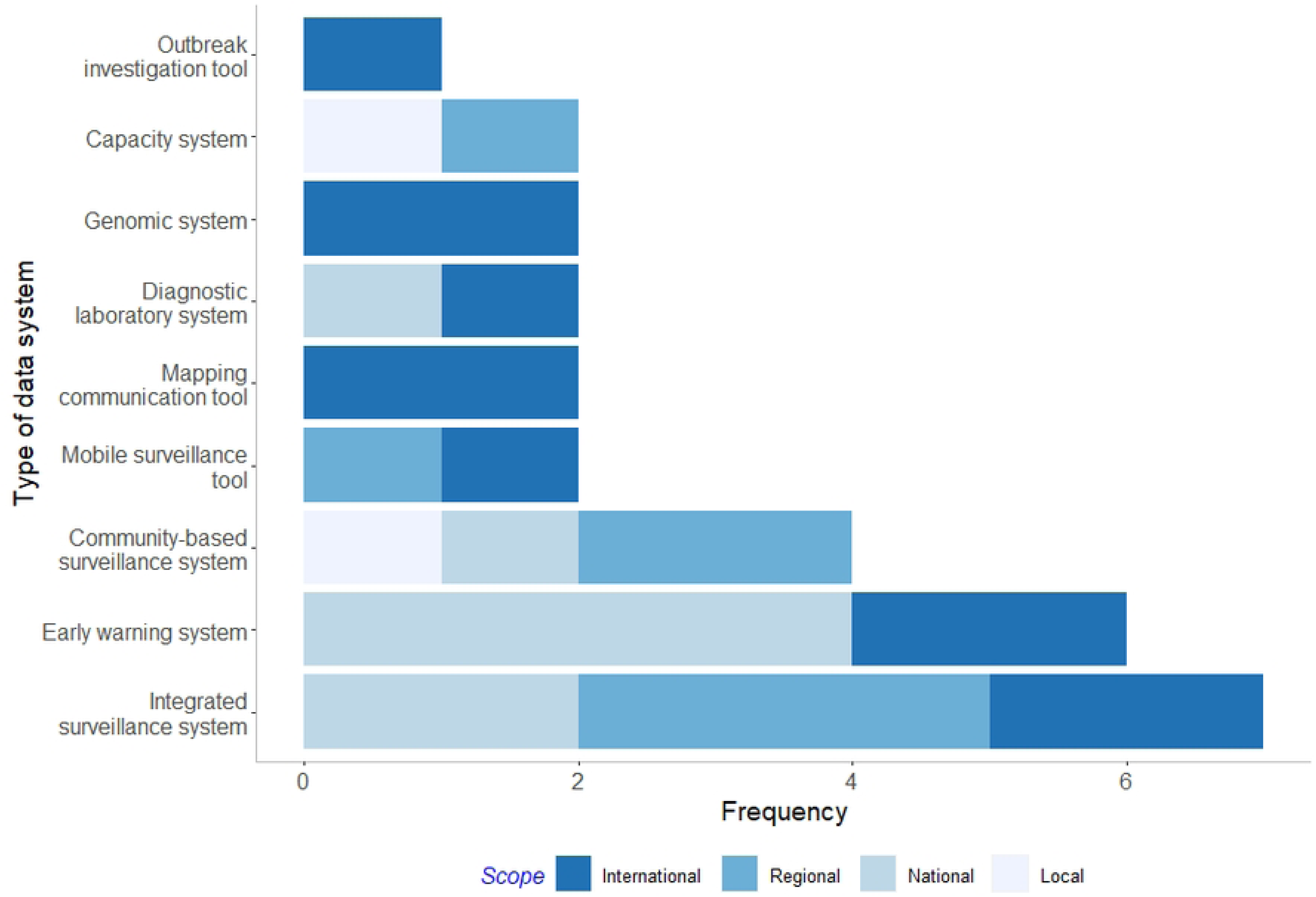
Frequency of Design data systems by type and scope.

The varying scopes of data systems demonstrate evidence of data sharing for CSID outbreak detection and response across local (36, 46), national (32, 38, 40, 43, 46, 47, 49), regional (33, 34, 39, 41, 44), and international levels (30, 31, 33, 35, 37, 42, 45, 48). For example, a local community-based surveillance system in forested areas of Guinea facilitated information exchange across human, animal, and environmental sectors for effective Ebola and Lassa fever response (36). At the national level, the piloted Kenya Animal Biosurveillance System was an early warning system aimed at timely data sharing of Rift Valley fever-associated risk factors and syndromes to detect livestock disease and respond before spill-over to humans (38). Regional data-sharing was evident across the Community-based Active Surveillance and District-based Passive Surveillance systems, part of the Southern Africa One Health Surveillance Strategy, which uses EpiCollect mobile software to enhance disease event detection in human and animal populations, and share data across prospective sectors (33). International structures included mapping communication tools such as MosquitoMap (45) and VizHub (30), which enable online sharing of maps that quantify risk of vector abundance or pathogen transmission with relevant stakeholders.

Design data system studies identified limitations in data availability, data sharing across sectors, and surveillance capacity. These limitations encompassed factors such as funding (34, 46) and data availability or reporting issues within the animal sector (38, 43). In the human sector, delays in reporting were often attributed to paper-based data transmission and inadequate infrastructure (33, 38, 40, 45). Collaborative efforts across the human, animal, and environmental sectors were hindered (36, 39, 41, 44, 47, 49) by the absence of standardised guidelines, policies, or frameworks (39, 44). Additionally, studies pointed out limitations in surveillance or laboratory capacity (45, 48, 49).

The identified evidence needs encompassed inclusion of vulnerable groups through digitally enabled participatory methods and increased cross-sectoral data sharing. Specifically, these needs encompass the integration of vulnerable groups into disease surveillance (33, 43, 48, 49) or control efforts (34) through adoption of participatory methods harnessing mobile phone technologies (38, 43). Additionally, studies advocated for incorporation of climate data into analyses to enhance understanding of CSID transmission and development of early warning systems (35), or long-term planning of response, control, and mitigation strategies (2). Furthermore, studies described a need for greater political will and capacity to facilitate data-sharing across human and animal sectors (31, 33, 41, 43, 48, 49) underpinned by established data standards, protocols, or centralised platforms (33, 38–41, 44, 45, 49), the pooling of multisectoral funds to support One Health policies (41) and increased private sector (46) or governmental (34) financial support. Other needs included the incorporation of fine-scale data collected across broader regions (50, 51), utilisation of modelling studies to direct surveillance resources (42, 47, 50), and bolstered veterinary (42, 49) and laboratory (40, 41) capacities to effectively address challenges posed by CSIDs.

#### Usage data system studies

Of the 33 multidisciplinary studies that integrated data from data systems (i.e., Usage studies), 12 were risk mapping studies (30, 32, 52–61), ten were association studies (50, 62–70), seven were both risk mapping and association studies, and four were phylogenetic analyses (31, 51, 71, 72) (Fig 9, S3 Table). We did not find any attribution studies. The different study types included varying domains across human, animal, and environmental data. For example, most association studies (n = 7) investigated the association of environmental variables on human response variables, such as that of ambient temperature exposure on COVID-19 transmission, whereas all phylogenetic analyses (n = 4) investigated spill-over between humans and animals.

**Fig 9.**
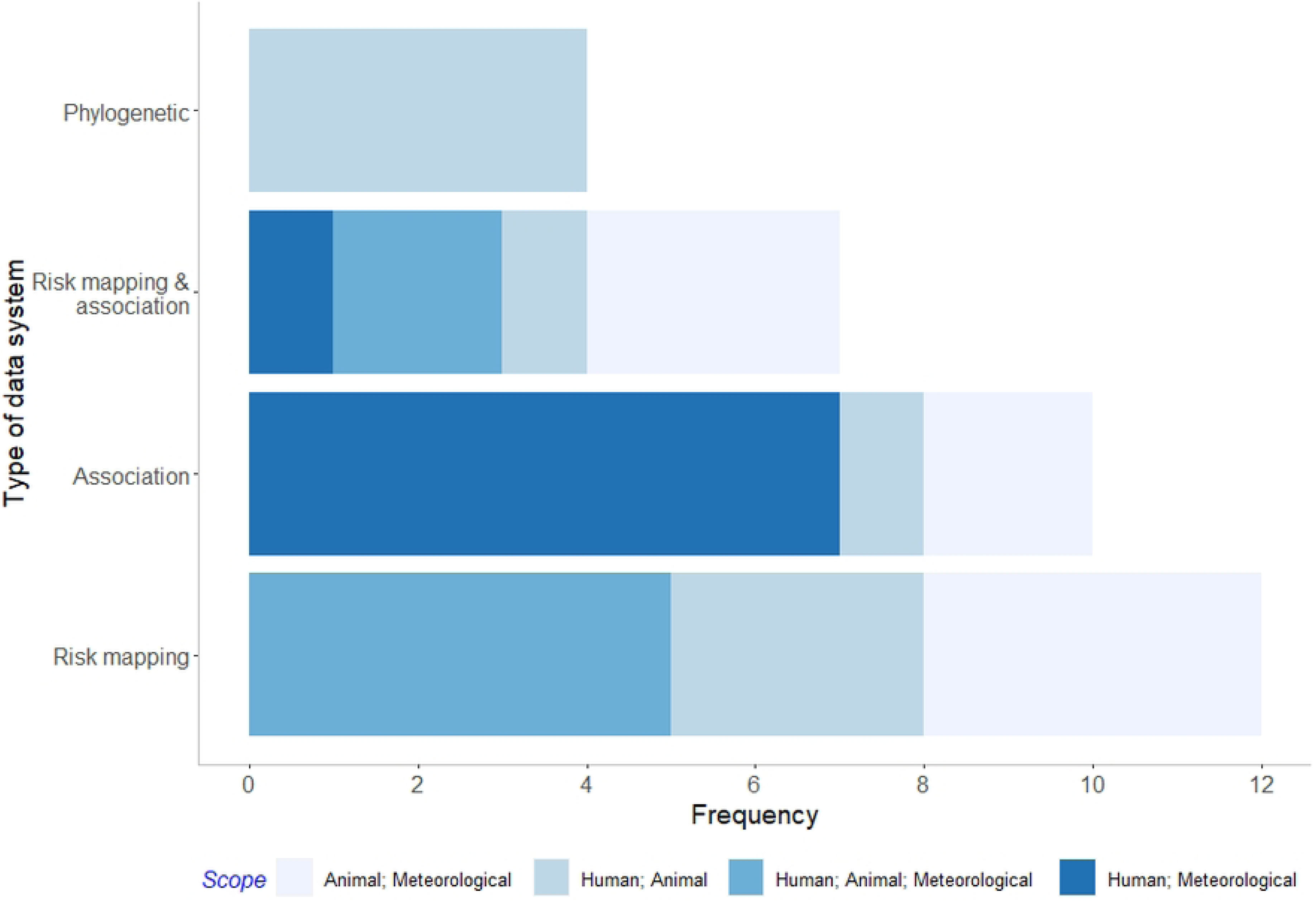
Frequency of Usage data systems study by study type and integration of data domain.

Evidence gaps identified in Usage studies surrounded data availability and cross-sector data sharing or coordination. Specifically, gaps revolved around inadequate availability of animal health data, often stemming from weak surveillance across the animal sector (52, 53, 65, 73, 74) or spatial biases in sampling and surveillance data (2, 51, 58, 71, 72). Additionally, deficiencies were noted in coordination between human, animal, and environmental surveillance systems (57) compounded by limitations in reporting capacity (31, 70) and limited temporal and spatial resolution of selected environmental data (75).

Evidence needs described in Usage studies included increased empirical data collection during outbreaks (52, 53) or routine surveillance (65) supported by participatory approaches to ensure data collection and inclusion of vulnerable groups (57). Furthermore, there was a call for the adoption of an integrated, One Health approach to surveillance, emphasising cross-sectoral and transboundary collaboration (31) alongside improved data sharing across animal and human sectors (61, 71, 72). Such initiatives should be supported by established data standards (62). Additionally, studies recognised the need for human and animal data linkage on a fine spatial scale (50–53), coupled with the integration of environmental data to deepen our understanding of the impact of climate change on CSID transmission (2, 58, 59, 64, 67, 76). These insights would inform the allocation of surveillance or control strategies (59, 63, 75, 77).

#### Gender-sensitivity of data system studies

Of the data system studies that included human data (n = 41), 38 studies were considered gender-blind, two were considered to be gender-sensitive (37, 40), and one gender-specific (36). Of the gender-sensitive studies, one described a system for predicting the causative pathogen of an outbreak which was programmed to disaggregate by gender and age (37), and one study reported Rift Valley fever cases by gender (40). The authors of the gender-specific study included women as key stakeholders in a surveillance data system, acknowledging their important role in disease detection despite their frequent under-representation (36).

While gender-blind studies did not consider gender, some considered other marginalised groups including rural communities (43, 73), socioeconomically marginalised groups (43, 49, 73), or youth groups (46). Notably, two studies placed an emphasis on understanding the association of multiple demographic variables, such as age and socioeconomic status, with COVID-19 transmission, but did not include gender as a variable in the analysis (70, 78).

### Narrative Synthesis: Economic studies

The 18 economic studies were comprised of economic evaluations (n = 10), which included CEAs and CUAs; costing studies (n = 3); and priority setting papers (n = 5).

#### Economic evaluation studies

Economic evaluation studies (n = 10) investigated the cost effectiveness of vaccination strategies (79–83), non-pharmaceutical interventions (NPIs) (84–86), or clinical critical care treatments (87, 88), and were limited to COVID-19 (n = 8) or Ebola (n = 2) (Table 2).

**Table 2:** Summary of economic evaluation studies.

| **First Author** | **Year of publication** | **Aims** | **CSID(s)** | **Population considered** | **Intervention/ comparators** | **Uncertainty analysis** | **Gender-lens** |
| --- | --- | --- | --- | --- | --- | --- | --- |
| Orangi (83) | 2022 | CEA to evaluate the potential epidemiological impact and cost-effectiveness of different vax roll-out scenarios in a Kenyan population that has already acquired a high-level immunity due to prior infections. | COVID-19 | Population of Kenya | COVID-19 vaccination roll-out strategies. | Univariate and probabilistic sensitivity analysis. | Gender-blind research |
| Reddy (84) | 2021 | CEA to assess clinical and economic outcomes and cost-effectiveness of COVID-19 control strategies in KwaZulu- | COVID-19 | Population of KwaZulu-Natal | Combinations of five NPIs across R = 1.5 and 1.2 epidemiological scenarios. | One-way and multiway sensitivity analysis | Gender-blind research |
|  |  | Natal province, South Africa. |  |  |  |  |  |
| Reddy (79) | 2021 | CEA to evaluate clinical outcomes and cost effectiveness of a COVID-19 vaccination program in South Africa. | COVID-19 | Population of South Africa | COVID-19 vax supply and vaccination pace strategies among $R_e = 1.4$ and 2-wave scenarios. | One-way and multi-way sensitivity analysis | Gender-blind research |
| Asamoah (85) | 2020 | CEA to evaluate NPIs against COVID-19 in Ghana and model transmission dynamics. | COVID-19 | COVID-19 cases in Ghana | Combinations of five NPIs. | Varied estimated parameter values to formulate parameters that would generate 'optimal control model' | Gender-blind research |
| Ruiz (80) | 2023 | CEA of three key vaccination policy questions in response to COVID-19 pandemic in Nigeria (which vax, who to prioritise, and how to deliver) | COVID-19 | Population of Nigeria | COVID-19 vax types, viral vectors, delivery method, prioritisation groups. | Analysis conducted over a range of scenarios and presented as main results | Gender-blind research |
| Liu (81) | 2022 | CEA investigating impacts of timing on COVID-19 disease burden in AU states. | COVID-19 | Population across 27 African countries | Vaccination (viral vector vaxs and mRNA vaxs), and no vaccination | Sensitivity analysis | Gender-blind research |
| Obeng-Kusi (82) | 2022 | CEA investigating value-based price for an Ebola vax package considering how prices compare across countries that have experienced outbreaks (Democratic Republic of Congo, Liberia, Sierra Leone, Uganda) | Ebola | 2014-2016 EVD cases in DRC, Liberia, Sierra Leone, Uganda. | Ebola vax package (vax, storage, maintenance, and administration) | Probabilistic sensitivity analyses | Gender-blind research |
| Kairu (87) | 2021 | CEA of investments in essential care and advanced critical care | COVID-19 | Hospitalised COVID-19 patients | Essential Care, Advanced Critical | One-way sensitivity analysis and a | Gender-blind research |
|  |  | during COVID-19 pandemic in Kenya. |  | admitted between March 2020 and January 2021. | Care, or maintaining status quo. | probabilistic sensitivity analysis. |  |
| Beshah (88) | 2023 | CEA of COVID-19 critical care intervention approaches: non-invasive (oxygen without intubation) and invasive (intubation) management in Ethiopia. | COVID-19 | COVID-19 patients (age 18+) enrolled at treatment centre and home-based isolation care between Jan 1 - May 31, 2021. | Non-invasive and invasive COVID-19 critical case management. | Probabilistic and one-way sensitivity analysis | Gender-blind research |
| Kellerborg (86) | 2020 | CEA of earlier interventions against Ebola in Sierra Leone. | Ebola | EVD patients from 2014-2016 Sierra | Time period of interventions taking place (original time or 4 weeks earlier) | Univariate sensitivity analysis | Gender-blind research |
|  |  |  |  | Leone<br>outbreak |  |  |  |

Abbreviations: CEA: Cost-effectiveness analysis; Vax: Vaccine.

Evidence gaps identified by the economic evaluation study authors highlighted significant deficiencies in data availability in relation to intervention efficacy and resource use/cost information and limited regional capacity for the conduct of economic evaluations. Specifically, studies reported a lack of data availability surrounding COVID-19 vaccination effectiveness (83), COVID-19 immunity data (79–81, 83), and costing data (83, 84). Some studies noted a lack of local data regarding COVID-19, necessitating reliance on UK estimates (83) or rendering the models unparameterizable to the local setting (87). Others discussed limited regional capacity for economic evaluations, specifically highlighting such aspects as an absence of established willingness to pay thresholds in South Africa (79) or more generally across LMICs (82); a limited ability to compare cost-effectiveness estimates due to a lack of economic evaluations (88); or a failure to consider health system restraints (80). Additionally, one study addressed the challenge of incorporating human productivity loss into economic analyses of Ebola (86).

Stated evidence needs included broadening cost-effectiveness analyses of vaccines to incorporate the far-reaching impacts of COVID-19 (83) and accounting for wealth inequity through subpopulation analyses (82). Additionally, extrapolation of epidemiological and economic models to other settings for informing vaccination purchasing (80), and incorporation of timing into evaluations for more realistic cost-effectiveness assessments (81).

#### Costing studies

The three costing studies analysed the costs of various initiatives across vaccination programs, a surveillance system, and outbreak response. One study presented cost estimates of a COVID-19 vaccination program in Ghana utilizing the COVID-19 Vaccination Introduction and Deployment Costing Tool, and undertaken under the leadership of the Ghana HTA program (89); another was a micro-costing study of a community-based surveillance (CBS) system for detecting Ebola and COVID-19 (among non-target CSIDs) in Sierra Leone (90); and the final study costed the US CDC’s response to Ebola outbreaks across Sierra Leone, Guinea, and Liberia (91) (Table 3).

**Table 3:** Summary of costing studies.

| **First Author**<br>**(Reference)** | **Year of**<br>**publication** | **Aims** | **CSID(s)** | **Population considered** | **Uncertainty**<br>**analysis** | **Gender-lens** |
| --- | --- | --- | --- | --- | --- | --- |
| Nonvignon<br>(89) | 2022 | Costing study to present cost estimates of COVID-19 vax program in Ghana utilising the COVID-19 Vax Introduction and Deployment Costing tool. | COVID-19 | All Ghanaians aged 16 years and above and are not pregnant. | Multi-way sensitivity analysis. | Gender-blind research |
| Mergenthaler<br>(90) | 2023 | Costing study of CBS in Sierra Leone as part of the electronic IDSR systems in context of Ebola and COVID-19. | Ebola;<br>COVID-19 | Sierra Leone health system (Micro-costing study from a health system perspective) | NR | Gender-blind research |
| Carias (91) | 2018 | Costing study of the US CDC's response to Ebola outbreaks in Sierra Leone, Guinea, and Liberia. | Ebola | Three Ebola outbreak clusters from 2016 in Sierra Leone, Guinea, and Liberia/Somalia. | NR | Gender-blind research |

**Table 4:** Summary of priority setting studies.

| **First Author** | **Year of publication** | **Aims** | **CSID(s)** | **Relevance to priority setting** | **Gender-lens** |
| --- | --- | --- | --- | --- | --- |
| Anantha-krishnan (92) | 2022 | Discuss examples of LMICs using HTAs during COVID-19 response and discuss broader HTA. | COVID-19 | Advocates for HTA use in pandemic response through case studies across LMICs. | Gender-specific research |
| Mosam (94) | 2020 | Discuss economic evaluation, community participation, and ethical considerations in priority setting of COVID-19 response in South Africa. | COVID-19 | Advocates for development of reliable but rapid processes for resource allocation during pandemic preparedness, highlighting importance of economic evaluation. | Gender-blind research |
| Kapiriri (95) | 2022 | Discuss how priority setting and resource allocation could be integrated into WHO pandemic preparedness framework to inform COVID-19 pandemic response, with Ebola examples. | Ebola;<br>COVID-19 | Provides a framework for incorporating economics across the WHO pandemic preparedness stages (pre-epidemic preparedness, alert phase, control phase, evaluation phase). | Gender-specific research |
| McCaul (93) | 2022 | Discuss COVID-19 Evidence Network to support Decision-making (COVID- | COVID-19 | Describes COVID-END, a global initiative of 50 evidence synthesis or support groups | Gender-blind research |
|  |  | END) body utilising South Africa case study. |  | for generation and communication of trustworthy, rapid, and equitable evidence syntheses to inform clinical and public health decisions and vaccination rollouts. |  |
| Zinsstag<br>(29) | 2016 | Address options for cost-effective control of animal disease and zoonoses in pastoral areas, as well as for disease surveillance and the financing of animal health services, utilising case studies in Ethiopia and Eastern/Western Africa. | Influenza;<br>Ebola; Rift<br>Valley fever | Discusses frameworks for evaluating economic efficiency of animal disease control across stages of pandemic preparedness, with a particular focus on surveillance. | Gender-blind<br>research |

Abbreviations: HTA: Health Technology Assessment

Evidence gaps described in the studies included delays in reporting by community health workers within the CBS and inconsistency in performance incentives being distributed (90), and lack of cost records attained within Ebola outbreaks resulting in the potential underestimate of costs (91).

Evidence needs included using the established COVID-19 Vaccine Introduction and Deployment Costing (CVIC) tool for costing the introduction of vaccinations across LMICs (89) and incorporating reporting of symptoms of COVID-19 and other future emerging infections into existing CBS for an early-warning systems (90).

#### Priority setting studies

Priority setting papers described the role of economic evaluations and associated priority setting tools or guidelines to support decision-making. Among the five studies, one described the application of HTAs for equitable decision-making in COVID-19 in LMICs (92), while another described the Evidence-to-Decision Framework utilised by the South African Grading of Recommendations Assessment, Development and Evaluation (GRADE) network to generate 42 rapid reviews for timely COVID-19 response (93). Another study considered economic evaluation, community participation, and ethical considerations in priority setting for COVID-19 response (94), and one discussed economic evaluation and evidence synthesis processes for priority setting across the four stages of pandemic preparedness (95). Lastly, a study discussed potentially cost-effective control strategies for animal and zoonotic diseases, including Rift Valley fever, Influenza and Ebola, in pastoralist populations (29), marking the sole economic study to address CSIDs beyond Ebola or COVID-19.

The authors of priority setting studies highlighted several evidence gaps, including potential barriers to the application of HTAs and challenges stemming from insufficient collaboration or capacity within the animal sector. Specifically, these gaps encompassed the complexity of HTAs as a barrier to decision-making (92, 95); a lack of collaboration resulting in duplicated efforts and resource wastage during the COVID-19 pandemic (93); limitations in animal disease surveillance and data maintenance impeding timely outbreak detection; and the absence of a stringent framework for monitoring the success of animal interventions leading to hesitancy in investment by institutional donors (29).

In addition, authors highlighted the need to increase regional capacity and collaboration while utilising participatory methods to enhance economic benefits and achieve equitable resource allocation. This would include leveraging strong leadership with political and institutional support to integrate HTAs into policy-making and foster regional technical expertise (92) and facilitating collaboration between designated working groups to prevent duplication of evidence synthesis efforts (30). Additionally, studies suggested employing mobile phone technologies in near real-time community-based surveillance systems, integrating multi-disease zoonotic CSID control approaches to optimise intervention benefit-cost ratios in pastoral areas (29), and enhancing community participation and expert ethical consideration for equitable resource allocation during pandemics (94, 95).

#### Gender-sensitivity of economic studies

Of the economic studies, 16 were considered gender-blind according to the WHO intersectionality toolkit (10), and two were gender-specific. Of the gender-specific studies, one study discussed gender and socioeconomically marginalised groups as stakeholders that could benefit from HTAs, noting that power dynamics between groups could limit their inclusion into HTA processes (92). The other gender-specific study discussed how vulnerability to pandemics could be exacerbated by gender and socioeconomic status, highlighting the importance of their consideration into research priorities and resource allocation within pandemic preparedness (95).

Of the gender -blind studies, two considered other vulnerable groups in the analysis, focusing on the elderly as high risk to COVID-19 (83) or implementing cost-effective control strategies against zoonoses in pastoralist communities (29).

### Critical appraisal within sources of evidence

A critical appraisal of evidence was not conducted.

## Discussion

We conducted a rapid scoping review to identify research needs for improving priority setting for pandemic preparedness in sub-Saharan Africa, focusing on data system and economic studies related to 14 CSIDs with pandemic potential. Our analysis revealed that while African author involvement is increasing, there remain few studies led by senior authors from African institutions, underscoring the need for research funders to support more locally led research efforts. Peer-reviewed economic studies on CSIDs are limited, with most research focusing on COVID-19 and Ebola and originating from South Africa, highlighting a gap in resource allocation and capacity for other diseases. The high degree of heterogeneity across data system studies, along with poor data sharing across siloed sectors, indicates a need for clear data system definitions and guidelines to develop interoperable and scalable solutions. Additionally, the limited inclusion of gender-sensitive perspectives in both economic and data system studies points to a significant gap that should be addressed to ensure more comprehensive and equitable pandemic preparedness strategies. Taken together, these identified gaps present important opportunities to enhance priority setting, improve resource allocation, and achieve more equitable health outcomes across the region.

### Gaps and opportunities

Studies with all authors affiliated with African institutions were limited throughout the study period, with the first such study appearing only in 2018. Additionally, the majority of first authors (69%) and last authors (63%) were affiliated with institutions outside of Africa. Although our sample size is small, these trends are consistent with recent research findings on climate change research capacity within the East African region (96). These regional gaps may indicate a lack of local capacity in data systems or economic studies. However, they could also reflect global power dynamics and funding inequalities, where international funders predominantly support global institutions. This often results in first and last authors being from outside Africa, potentially leading to research questions and priorities being set without adequate input from African researchers. Addressing these disparities is important for fostering a more equitable and effective research environment.

The concentration of economic studies across COVID-19 and Ebola, and primarily conducted in South Africa, suggests a broader deficiency in capacity and technical expertise for conducting similar evaluations across the rest of the WHO AFRO region and for other CSIDs. This observation is consistent with findings from a previous review that assessed HTA institutionalisation in sub-Saharan Africa (97), which highlighted a lack of expertise and tools for context-specific decision-making aligned with locally agreed frameworks for economic evaluation.

To address these deficiencies, developing national HTA frameworks that include the use of context specific economic evaluations could significantly enhance priority-setting systems (17, 98). These frameworks should incorporate a diverse range of stakeholder perspectives and extend the usual role of HTAs to address key questions, such as how to fund or create priority-setting processes that link climate change and CSID response, and what existing structures support intersectoral actions for pandemic preparedness. Such country-specific HTA frameworks would help assess existing resources and could be applied to a range of CSIDs beyond COVID-19 and Ebola, including emerging threats such as Mpox, which was recently declared a PHEIC by the WHO (99).

Additionally, HTA-based priority setting approaches and economic evaluations could be applied to earlier stages of the pandemic preparedness framework, to assess prevention and detection strategies. The prominence of COVID-19 studies likely accounts for most of the identified peer reviewed economic studies focusing on later stages of pandemic preparedness, due to the epidemiological situation at the time of publication. Economic evaluations targeting earlier stages, for example on preventing emerging infectious diseases with pandemic potential, could highlight the benefits of investing in prevention and support the use of financing tools like Development Impact Bonds, where donors share the risk of investing in high-risk prevention strategies (29).

We found a large degree of heterogeneity in data system literature, reflecting a lack of clear and universally agreed-upon definitions of data systems related to pandemic preparedness. Establishing these definitions would guide the creation of more effective data systems that provide decision-makers with accurate and timely information. For pandemic preparedness against CSIDs, these definitions should emphasise interoperability and data harmonisation to enable the routine integration of diverse data sources, such as human health, animal health, and environmental data.

Such data systems would enhance data sharing across traditionally siloed sectors, addressing a significant gap identified by both data system and economic studies in this review. Sectoral silos and a lack of digital integration platforms are common barriers to data sharing, which hinder evidence-driven decision-making in a One Health context (100, 101), despite recommendations from the One Health Joint Plan of Action (101). The Digital One Health (DOH) framework, based on the FAIR principles (Findability, Accessibility, Interoperability, and Re-use), aims to address these barriers by consolidating data-sharing across five key pillars (102). These pillars focus on harmonising and automating standardisation according to ethical and legal guidelines, determining which data can be shared, such as pathogen characteristics and patient gender, or withheld, such as patient name or contact. The framework is currently being piloted in antimicrobial resistance surveillance in Uganda (100). Tailoring such frameworks to the specific challenges faced across decision-making in pandemic preparedness could bridge siloed efforts, enhance coordination, and provide high-quality data for pandemic preparedness decision-making.

Limited gender-sensitivity across both data systems and economic studies may result in inadequate consideration of gender-specific impacts and needs in pandemic preparedness, despite a growing body of evidence that highlights the differential effects of pandemics on various genders (7–10). Participatory approaches can provide methodologies for gender-transformative change, by identifying vulnerable groups that are context specific and including them into research and subsequent decision-making (10). Digital participatory methods, such as use of mobile phone applications for reporting animal cases can facilitate near-real-time reporting of suspected outbreaks (103), and also help to incorporate hard-to-reach populations into decision-making processes. Importantly, digital inequities may hinder the inclusion of marginalised groups, particularly women and girls (9, 103), due to limited access to technological education (104), especially in low- and middle-income countries. Therefore, developing participatory digital tools that are culturally and contextually informed and investing in digital skills for those who need them most, should be an important consideration for pandemic preparedness.

We did not find any data system studies that directly attributed human CSID-related health outcomes to human-caused climate change, despite numerous studies assessing associations between environmental variables and health outcomes. This reflects a broader gap in attribution research; a recent report identified only 13 studies globally since 2013 that rigorously assessed health impacts attributable to human-caused climate change (105), with the only CSID study that attributed human-caused climate change to increased childhood malaria in sub-Saharan Africa (106). The lack of attribution research on CSIDs is notable, given that the impact of climate change on CSID-related health outcomes is a priority for attribution research (105). Enhancing data integration across human, animal, and environmental domains through interoperable systems could facilitate assessments of long-term trends and associations, inform counterfactual analyses of climate scenarios, and support economic evaluations of mitigation and adaptation strategies (107). Such efforts could significantly strengthen the evidence base on the health impacts of climate change on CSIDs.

#### Limitations

We did not conduct a grey literature search due to the expedited timelines of the rapid scoping review, likely omitting relevant literature including governmental policy reports that may describe governmental data-system and economic evaluation use. To mitigate this limitation, we utilised the expertise of the project steering committee and their experience across complementary reviews, screening additional studies they identified as potentially relevant. Secondly, we only included papers published in English. As a result, an English-language bias may have caused the omission of studies published by authors from countries where English is not the first language, the inclusion of which may alter the conclusions drawn. Thirdly, to manage the number of publications, we only included studies that explicitly mentioned target CSIDs. This may have excluded some publications relevant to pandemic preparedness of CSIDs in less explicit yet important ways, such as focusing on the mode of transmission rather than the pathogen or illness. Broadening the scope to include these studies may provide useful insights into wider data systems or economic evaluations for pandemic preparedness, such as the use of Natural Capital Accounting to assess the broader impacts of disease outbreaks on ecosystem services and public health, which would be excluded unless it explicitly mentioned a target CSID. Finally, the rapid nature of the review process precluded in-depth quality assessments of the included studies. Future reviews should incorporate comprehensive quality appraisals to better understand the reliability and validity of the evidence base.

## Conclusions

Our rapid scoping review revealed significant evidence gaps in data systems and economics for priority setting of pandemic preparedness in sub-Saharan Africa. Key issues include limited African-led research, a scarcity of peer-reviewed economic studies on most CSIDs, inadequate data sharing across human, animal, and environmental domains, and insufficient application of gender-sensitive perspectives. Addressing these gaps presents important opportunities to enhance decision-making in pandemic preparedness and ensure more effective and inclusive responses to emerging infectious disease threats. This need is especially urgent given the changing climate, which increases the risk of CSID pandemics, as demonstrated by the recent Mpox outbreaks.

## Data Availability

The data is available at the Open Science Framework at https://osf.io/fn8rw/?view_only=5961c57df32741b884fc2181a0da4ceb

## Acknowledgements

This work was supported by the International Development Research Centre (PO5001440). The funders had no role in study design, data collection and analysis, decision to publish, or preparation of this manuscript. The opinions expressed here belong to the authors, and do not necessarily reflect those of the funder.

## CRediT author statement

**Ellie A. Delight:** Methodology; Formal analysis; Investigation; Data Curation; Writing - Original Draft; Writing - Review & Editing; Visualization. **Ariel A. Brunn:** Conceptualization; Methodology; Validation; Investigation; Resources; Writing - Original Draft; Writing - Review & Editing; Supervision; Project administration; Funding acquisition. **Francis Ruiz:** Conceptualization; Methodology; Investigation; Resources; Writing - Original Draft; Writing - Review & Editing. **Jessica Gerard:** Formal analysis; Investigation; Data Curation; Writing - Original Draft; Writing - Review & Editing; Visualization. **Jane Falconer**: Methodology; Software; Validation; Investigation; Data Curation; Writing - Review & Editing. **Yang Liu**: Conceptualization; Investigation; Writing - Review & Editing. **Bubacarr Bah:** Conceptualization; Investigation; Writing - Review & Editing. **Bernard Bett:** Conceptualization; Investigation; Writing - Review & Editing. **Benjamin Uzochukwu:** Conceptualization; Investigation; Writing - Review & Editing. **Oladeji K. Oloko:** Conceptualization; Writing - Review & Editing. **Esther Njuguna:** Conceptualization; Writing - Review & Editing. **Kris A. Murray**: Conceptualization; Methodology; Validation; Investigation; Writing - Review & Editing; Supervision; Project administration; Funding acquisition.

## Competing interests

FR was supported by funding received from the Bill and Melinda Gates Foundation (OPP1202541) through the International Decision Support Initiative (iDSI). All other authors declare that they have no actual or potential competing interests that could have influenced the work reported herein.

## Supporting information

**S1 Table: Screening guidance**

**S2 Table: Summary of Design Data system studies**

**S3 Table: Summary of Usage Data System studies**

## Notes

### Clinical Protocols

https://doi.org/10.17605/OSF.IO/VTGMC

### Funding Statement

This work was supported by the International Development Research Centre (PO5001440). The study was conducted independently from the International Development Research Centre.

### Author Declarations

This study is a rapid scoping review, guided by PRISMA-ScR and did not involve any primary data collection or human participants. Therefore, ethical approval was not required. All data analysed in this review were obtained from publicly available, peer-reviewed publications.

